# The effect of physical activity timing on insomnia and sleep quality: a randomized cross-over trial in older adults

**DOI:** 10.64898/2026.05.18.26353463

**Authors:** Gali Albalak, Raymond Noordam, Marjan van der Elst, Thomas Drop, Eloy Caneda Cabrera, Luuk Oudendijk, Gert Jan Lammers, Marijke Gordijn, Laura Kervezee, Vasileios Exadaktylos, David van Bodegom, Diana van Heemst

**Affiliations:** Department of Internal Medicine, Section of Gerontology and Geriatrics, Leiden University Medical Center, PO Box 9600, Albinusdreef 2, Leiden, 2300 RC, The Netherlands; Department of Clinical Epidemiology, Leiden University Medical Center, Leiden, the Netherlands; Health Campus the Hague/Public Health and Primary Care, Leiden University Medical Center, the Hague, the Netherlands; Department of Neurology, Leiden University Medical Centre, Leiden, the Netherlands; Stichting Epilepsie Instellingen Nederland, Sleep-Wake Centre, Heemstede, the Netherlands; Chrono@Work, Groningen, the Netherlands; Chronobiology Unit, Groningen Institute for Evolutionary Life Sciences, University of Groningen, Groningen; Group of Circadian Medicine, Department of Cell and Chemical Biology, Leiden University Medical Center, PO Box 9600, Einthovenweg 20, 2333 ZC, Leiden, The Netherlands; Centre for Human Drug Research, Leiden, Netherlands; Leyden Academy on Vitality and Ageing, Rijnsburgerweg 10, Leiden, 2333 AA, The Netherlands; Department of Public Health and Primary Care, Leiden University Medical Center, PO Box 9600, Albinusdreef 2, Leiden, 2300 RC, The Netherlands

## Abstract

**Background:** Insomnia symptoms are common in older adults. While observational studies suggest physical activity (PA) timing affects health outcomes, its effect on sleep remains unclear. We compared morning versus evening PA effects on insomnia severity and sleep quality in older adults with insomnia symptoms.

**Methods:** Eligible participants were aged 60–80 years with (sub)clinical insomnia (Insomnia Severity Index [ISI] score ≥10). In a randomized cross-over trial, participants engaged in coached PA in the morning (10:00–11:00) or evening (19:30–20:30) for 14 days each. ISI scores were assessed post-intervention. Objective sleep parameters—duration, latency, efficiency, and timing—were assessed with a Withings Sleep Analyzer under the mattress. Subjective sleep quality was reported daily via smartphone app. Salivary dim light melatonin onset (DLMO) was measured on the final day of each intervention.

**Results:** Of 37 participants (mean ISI 14.3 ± 3.3), 27 completed the study (mean age 69.8 ± 5; 63% women). ISI scores improved after both morning (Δ -2.5; 95% CI: -1.14, -3.83) and evening (Δ -2.0; 95% CI: -0.63, -3.38) activity relative to baseline, but were not different between interventions. Compared to evening activity, sleep midpoint occurred earlier with morning activity (03:40 vs 04:00; Δ -20 min; 95% CI: -31, -8). No differences in subjective sleep quality or DLMO were found. Exploratory analyses suggested insomnia scores improved specifically in late chronotypes following morning activity.

**Conclusions:** While morning vs. evening PA timing did not impact most sleep quality measures, it influenced sleep timing. Larger studies are needed to define optimal and personalized PA timing for improving sleep.

## Introduction

Poor sleep is an important risk factor for many (age-related) diseases. For example, extreme short or long sleep duration and insomnia have been associated with multiple age-related chronic diseases such as diabetes, obesity, dementia, cardiovascular disease and cancer,(1, 2) with evidence for causality for multiple diseases.(3-5) The prevalence of sleep complaints increases with advancing age, and is more frequent among women compared to men.(6, 7) In the Netherlands, approximately 28% of the people aged 75 years and older have self-reported sleep problems.(8) Poor sleep can be caused or worsened by disturbances in the circadian clock that orchestrates and synchronizes internal processes and coordinates physiological, mental and behavioral pattens within a cycle of approximately 24 hours.(9) Shift work, the 24/7 society, social jetlag and aging may lead to circadian misalignment and poorer sleep quality.(10-12)

It has been shown that physical activity can synchronize circadian rhythms with each other and the external environment.(13) In a recent study with day active rodents, researchers found that daytime physical activity directly influences clock neuron activation and thereby strengthens the output of the clock.(14) Multiple observational studies conducted in large cohorts have shown associations between physical activity at a specific time of day (i.e., “chronoactivity”(15)) with outcomes such as metabolic health, cardiovascular health, mental health and even all-cause mortality.(15-21) Moreover, physical activity can cause a phase advance or phase delay in the sleep-wake cycle and other circadian rhythms depending on its timing.(22, 23) Yet, despite the increasing interest and mounting evidence for the relevance of chronoactivity(16, 21, 24, 25), the optimal circadian timing of physical activity for circadian health remains unclear and little is known about the influence of chronoactivity on sleep in older adults who frequently suffer from sleep problems.(8, 26) Moreover, as most research in this domain has focused on observational research embedded in prospective cohort studies, the direct effect of activity timing on health remains to be determined.

Considering the negative effects of circadian misalignment, the large number of older people suffering from sleep problems, and the seeming relevance of chronoactivity, we aimed to study the effects of timed physical activity on insomnia severity and other sleep-related outcomes in older adults with self-reported sleep problems included in the Older Adults Exercising ON TIME study.(27)

## Materials and methods

### Study setting and design

A detailed description of the design and rationale of this study has been published previously.(27) In short, the ON TIME study was a randomized cross-over intervention study conducted in the Leiden University Medical Center (LUMC, the Netherlands) between April and June 2024 (one month after the transition to daylight saving time). This study had two-arms and a superiority framework with an intervention allocation ratio of 1:1. It consisted of an eight-week protocol including a baseline visit, and three interventions that all participants followed consecutively and simultaneously in two randomly allocated groups, with a follow-up visit after each of the three interventions. The study and data collection largely took place at the participants’ home except for the training sessions that took place at an outdoor sports facility on LUMC grounds and the study site visits in the LUMC. The study protocol was approved by the Medical Research Ethical Committee of Leiden, The Hague, Delft (MREC-LDD), The Netherlands in June 2023 and can be found in the Clinicaltrials.gov register with protocol ID: NCT06613958 or in the Dutch register with protocol ID: NL82335.058.22/ (Centrale Commissie voor Mensgebonden Onderzoek, CCMO) https://onderzoekmetmensen.nl/nl/trial/56414.

### Participants

For this study, we included overall healthy older adults (between 60 to 80 years old) with long-lasting presence of (sub)clinical insomnia symptoms. Insomnia symptoms were defined by the Insomnia Severity Index (ISI).(28) Power calculations were done to define the sample size.(27) An ISI cut-off of 10 points was used as this was found to be optimal for identifying insomnia cases in a community sample.(28) Additionally, participants were asked if their sleep problems lasted three months or longer. **Supplementary table 1** shows an overview of all the in- and exclusion criteria. Participants were recruited through advertisements in flyers, (local) newspapers, social media, websites and local radio. For practical reasons, participants were only eligible for participation when they were from the municipality of Leiden or from neighbouring villages.

### Exposure

The ON TIME study consisted of three intervention periods; a sedentary period, an active morning period, and an active evening period, each lasting two weeks. During the sedentary period, participants were asked not to perform any exercise and minimize strenuous household-related activities. During the two active periods, participants were asked to perform moderate-to-vigorous physical activity every day either in the morning (from 10:00 to 11:00) or evening (19:30 to 20:30). This was done four times a week in a group setting guided by a trained professional and three times a week individually at a participant’s place of choice (e.g., home). With the exception of the designated exercise moment, participants were instructed to only engage in light physical activities throughout the day. A detailed description of the intervention can be found in the ON TIME study protocol paper.(27) Both groups started with the sedentary period of 14 days that served as a control or baseline period. Depending on the intervention arm allocated via randomization, participants subsequently either underwent the active morning intervention or active evening intervention first, followed by the remaining intervention. There were two washout periods of seven days, one between the sedentary period and first intervention and one between the first and second intervention to prevent or minimize potential carryover effects. Verification of the validity of the intervention was done by calculating hourly means of step count from the wrist-worn wearable (Cardiowatch 287-2, Corsano Health, The Hague, The Netherlands) and visual inspection of the line plots.

### Outcomes

#### Insomnia severity

The primary outcome of this study was insomnia severity measured by the Insomnia Severity Index (ISI). The ISI is a subjective measure of sleep quality and severity of insomnia complaints. It is a 7-item questionnaire (0-28 points; a score of 0-4 for each item) that is easy to use and has been validated as a screening tool and an outcome measure for sleep problems.(28, 29) Based on the ISI questionnaire, participants can be categorised as having: ‘No clinically significant insomnia (0-7 points)’, ‘Subthreshold insomnia (8-14 points)’, ‘Moderate severity clinical insomnia (15-21 point), and ‘Severe clinical insomnia (22-28 points)’. This questionnaire was conducted at baseline and on the 13^th^ day of each intervention period during the study visit.

#### Sleep quality

Multiple objective components of sleep quality were assessed with the Withings Sleep Analyzer(30, 31) (Withings, Issy-les-Moulineaux, France) that was installed under the participants’ matrasses before the start of the study by a trained research nurse or staff member. These sleep quality components included:

- Sleep duration: total minutes asleep.
- Sleep latency: total minutes of lying in bed before falling asleep.
- Sleep efficiency: ratio of total sleep duration to time spent in bed.
- Midpoint of sleep: the middle clock time point between sleep onset time and wakeup time.
- Number of awakenings between sleep onset and wake time.
- Minutes and percentage spent in sleep stages: minutes spent in light sleep, deep sleep and Rapid Eye Movement (REM) sleep.

Moreover, as a measure of perceived sleep quality, participants were asked to score the following statements (presented to the participants in the Dutch language) each morning with a visual analogue scale (0-100, 0=totally disagree with statement and 100=totally agree with statement):

- “I feel rested”.
- “I am satisfied with my sleep last night”.

These statements were presented to the participants as part of the ecological momentary assessment (EMA) with the mPath application (KU Leuven, Belgium).(32) Among baseline collected characteristics(27) the Munich Chronotype Questionnaire (MCTQ) was conducted at baseline to assess participants’ chronotype by determining the mid-point of sleep on free days corrected for sleep debt (MSF_SC_).(33)

#### Dim Light Melatonin Onset

On the last day of each intervention period (3 times in total), participants collected hourly saliva samples with 7 cotton swabs in a time range of 5 hours prior to bedtime to 1 hour after bedtime to assess Dim Light Melatonin Onset (DLMO), a commonly used marker of the phase of the circadian pacemaker.(34) Saliva samples were collected using Salivette (Sarstedt Ltd., Nümbrecht, Germany). Participants were instructed to stay at home in dim light conditions (with closed curtains) throughout the evening, starting one hour prior to the first measurement. Watching television was allowed when wearing sunglasses and participants were allowed to read by a dim screen light. Participants were asked to note when they took any medication. The collection of saliva was carried out in a sitting or supine position and participants were asked not to change position 5 minutes before performing the sampling. Participants were allowed to eat and drink 30 minutes after saliva was collected, with the exception of coffee, tea, drinks containing alcohol, chocolate, bananas and food and drink containing colorants. Additionally, participants were asked not to use toothpaste and any substances containing tetrahydrocannabinol (THC). Participants were asked to note any discrepancies to the protocol on the report form and to store the Salivettes in the refrigerator overnight. The Salivettes were collected, centrifuged and the saliva samples were then stored at -70 °C by the study team within 36 hours after collection. After completion of the ON TIME study, the saliva samples were analysed by Chrono@Work (Groningen, the Netherlands). To assess melatonin concentration, a double-antibody radioimmunoassay (RIA) was performed with an intra-assay variation coefficients of 32.5% at a mean concentration 1.0 pg/mL and 10.9% at a mean concentration 13.9 pg/mL for low and high concentration controls, respectively and an interassay variation coefficients of 30.9% at a mean concentration 0.9 pg/mL and 8.7% at a mean concentration 15.3 pg/mL for low and high concentration controls, respectively) (Direct Saliva Melatonin kit; Novolytix GmbH, Witterswil, Switzerland). DLMO was marked as the first time when linear interpolated melatonin concentrations exceeded the 4 pg/mL threshold.(35) To identify the clock time of DLMO, two researchers scored all individual melatonin onset plots separately to determine between which two measurements the melatonin levels crossed the DLMO threshold (4 pg/mL). In case of disagreements, the final decision was made by a third independent assessor. Then, an interpolated line between the two measurements surrounding DLMO was used to determine the final clock time of DLMO.

#### Statistical analyses

Characteristics of the study population were calculated and are presented as numbers with percentages (%) for categorical data and as mean with standard deviation (SD) for normally distributed or medians with interquartile ranges (IQR) for non-normal continuous variables. To address the primary outcome, paired t-tests were performed to compare the means of the ISI between baseline and any of the three interventions, as well as for comparing two interventions. The same analyses were performed for all additional objective and subjective sleep quality variables and DLMO. Phase angle refers to the time difference (in hours) between DLMO and sleep onset.(36) Results from the paired t-tests are shown as mean differences between the interventions (delta) with 95% confidence interval and p-value. A two-sided p-value below 0.05 was considered statistically significant. Non-normal outcome data was log-transformed with natural logarithm to approximate a normal distribution. Subsequently, differences with accompanying confidence intervals were transformed back into percentage differences to facilitate clinical interpretation of the results. These results can be interpreted percentage differences. Data was analyzed per protocol.

As an additional and purely explorative analysis, participants were divided into three groups according to their ISI deltas calculated from the difference between active morning and active evening; 1) participants with a ≥2 point higher ISI score after the active morning compared to the active evening, 2) participants whose ISI score did not change more than two points, and 3) participants with a ≥2 point higher ISI score after the active evening compared to the active morning. For the two groups that had a change of ≥2 points (threshold chosen to match power calculations), referred to as ‘benefit groups’, we exploratively listed the mean values for all sleep quality variables and their delta between the active morning and active evening group.

## Results

### Participant characteristics

From January until April 2024, 183 people expressed interest in participating to the study **(figure 1)**. After several screening steps, 37 participants were enrolled and randomly allocated in the study. 2 participants dropped out before baseline because of personal reasons and a change in medication. Between the baseline and start of the intervention another 3 participants dropped out due to technical or commitment issues. A total of 32 participants started with the first intervention period after which 5 participants dropped out leaving a total of 27 participants with complete data collection after the 3 intervention periods. **Table 1** shows the participant characteristics for the total study population as well as per randomization group. Participants included in the analyses were on average 69.8 (SD: 5) years old. The majority (63%) were women and participants were overall healthy (no mental complaints or severe physical complaints). Chronotype as assessed with the MCTQ was 3:09 AM (SD: 1:19).

**Figure 1.**
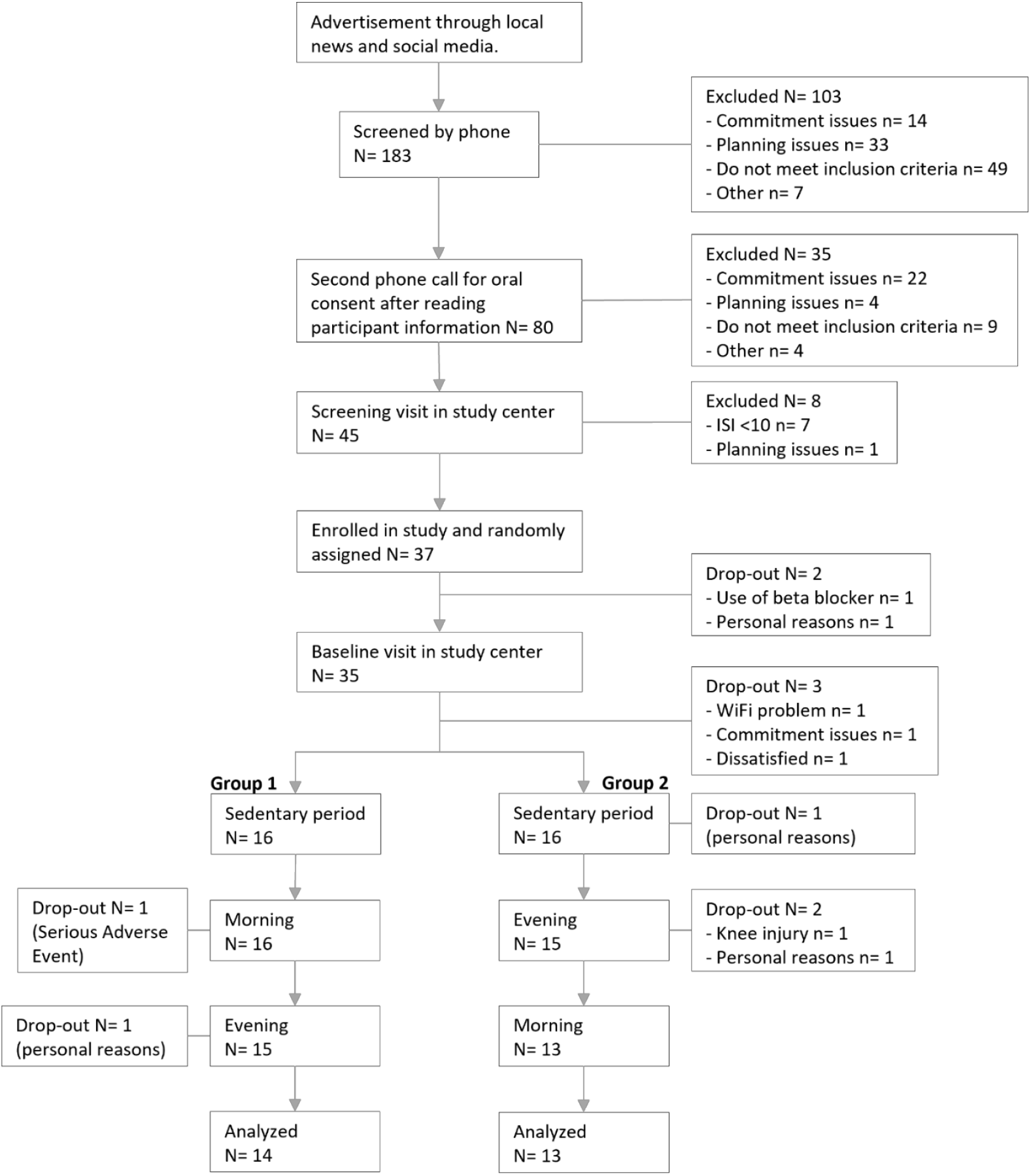
Flowchart showing all major screening and inclusion steps as well as the course of the study drop-outs.

**Table 1.** Baseline Participant characteristics.

|  | **Total population** | **Group 1\*** | **Group 2\*\*** |
| --- | --- | --- | --- |
| **N** | 27 | 14 | 13 |
| **Age**, mean (SD), years | 69.8 (5) | 69.1 (4.9) | 70.5 (5.3) |
| **Female sex**, No. (%) | 17 (63) | 8 (57.1) | 9 (69.2) |
| **BMI**, mean (SD), kg/m<sup>2</sup> | 27.8 (6.3) | 26.3 (4.5) | 29.5 (7.7) |
| **Chronotype** (h:mm) | 03:09 (1:19) | 3:05 (0:55) | 3:15 (1:50) |
| **Social jetlag** (min) | 0 (0-30) | 0 (0-53) | 0 (0-19) |
| **EQ-5D-3L**, median (IQR) | 0.81 (0.78-0.84) | 0.84 (0.81-1) | 0.81 (0.77-0.83) |
| **ADL**, median (IQR) | 0 (0-0) | 0 (0-0.25) | 0 (0-0.50) |
| **CCI**, median (IQR) | 0 (0-1) | 0 (0-1) | 1 (0-2) |
| **Baseline ISI**, mean (SD) | 14.3 (3.3) | 13.4 (3.9) | 15.3 (2.3) |
| **Years of education**, mean (SD) | 14.7 (4.6) | 15.6 (4.2) | 13.8 (4.9) |
| **Marital status** No. (%) |  |  |  |
| Unmarried | 4 (14.8) | 2 (14.3) | 2 (15.4) |
| Married | 15 (55.6) | 9 (64.3) | 6 (46.2) |
| widowed | 2 (7.4) | 1 (7.1) | 1 (7.7) |
| divorced | 6 (22.2) | 2 (14.3) | 4 (30.8) |
| **Smoking status**, No. (%) |  |  |  |
| Never | 9 (33.3) | 5 (35.7) | 4 (30.8) |
| Previous | 18 (66.7) | 9 (64.3) | 9 (69.2) |
| Current | 0 (0) | 0 (0) | 0 (0) |
| **Alcohol intake frequency** No. (%) |  |  |  |
| Never | 9 (33.3) | 2 (14.3) | 7 (53.8) |
| < 1 per week | 5 (18.5) | 3 (21.4) | 2 (15.4) |
| ≥ 1 per week | 13 (48.1) | 9 (64.3) | 4 (30.8) |
| **Hand grip strength**, mean (SD), kg |  |  |  |
| Left | 28.1 (9.7) | 29.4 (9.7) | 26.8 (9.9) |
| Right | 27.7 (9.6) | 29.4 (9.7) | 25.7 (9.5) |
| **Blood pressure**, mean (SD), mmHg |  |  |  |
| Systolic | 133.4 (16.9) | 132.9 (17.9) | 133.9 (16.5) |
| Diastolic | 80.7 (6.1) | 80.6 (5.8) | 80.9 (6.6) |
| **HbA1c**, mean (SD), mmol/mol | 40 (5.5) | 39.5 (6.5) | 40.5 (4.4) |

BMI, Body Mass Index; EQ-5D-3L, EuroQol Quality of Life 5 dimensions 3 levels; ADL, Activities of Daily Living; CCI, Charlson Comorbidity Index; ISI, Insomnia Severity Index; HbA1c, Hemoglobin a1c. Participants characteristics of the study population and per randomization group. Data is shown as number and percentage for categorical data or as mean and standard deviation for normally distributed data and median with interquartile range for non-normally distributed data. Social jetlag was defined as the difference (minutes) in midpoint of sleep on work-days and work free days. Chronotype was based on MSF\_sc ( sleep-corrected local time of mid-sleep on work-free days) and together with social jetlag calculated with data from the Munich Chronotype Questionnaire (MCTQ) assessed at baseline.
\* sedentary – active morning – active evening
\*\* sedentary – active evening – active morning

### Validation of the intervention

The mean daily step count was 2,748 ± 740, 3,079 ± 1,184, and 3,558 ± 1,154 steps for the sedentary, active morning and active evening period, respectively. A clear peak of activity was present for both the active morning and the active evening period whereas there was no clear peak visible during the sedentary period (**figure 2**) indicating that participants adhered to the prescribed timing of their physical activity.

**Figure 2.**
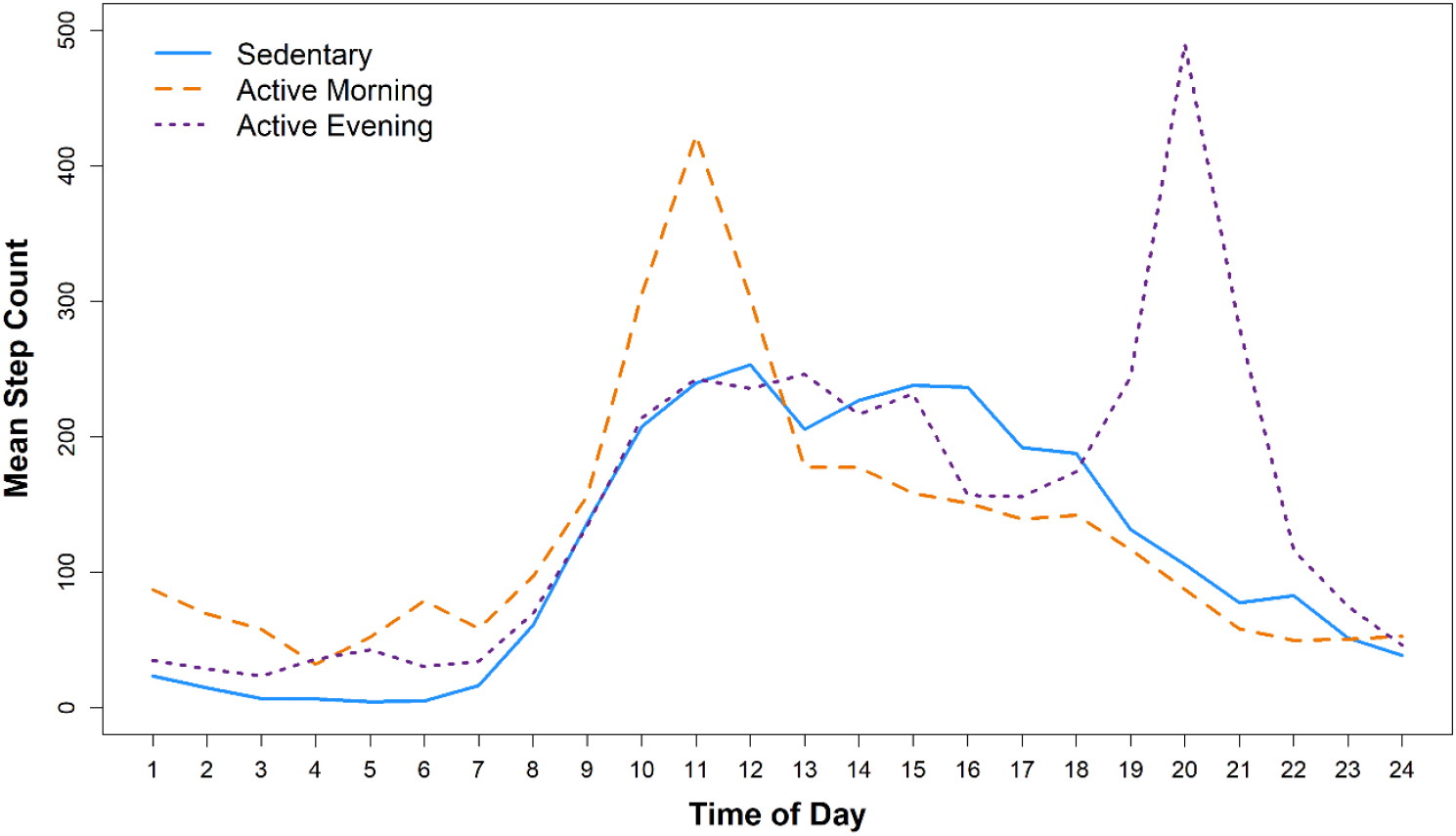
Line graph that displays the activity pattern per intervention period. Activity patterns were plotted based on hourly step counts as measured by the Cardiowatch 287-2 (Corsano Health, The Hague, The Netherlands) and based on average hourly step count of all seven days of the measurement week of all three intervention. The X-axis represents clock time and the Y-axis shows the hourly step count. The blue line depicts the activity pattern during the sedentary period in which no MVPA activities were undertaken. The orange line shows the activity pattern of the active morning intervention and the purple line represents the active evening intervention period.

#### Changes in insomnia severity

Insomnia Severity Index scores decreased after the sedentary, active morning, and active evening period compared to the baseline measurement (-1.9 points; 95%CI 0.77, 3.1, -2.5 points; 95%CI 1.14, 3.83, - 2.00 points; 95%CI 0.63, 3.38, respectively; **figure 3 and supplementary table 2**). There were no significant differences between the ISI scores of the intervention periods.

**Figure 3.**
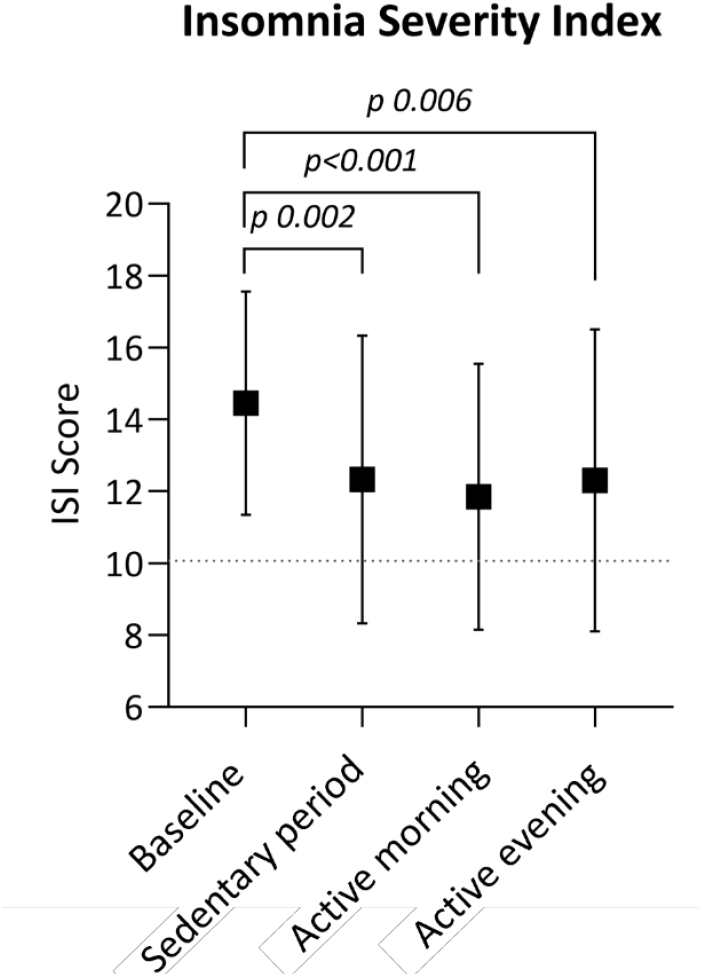
Figure shows results from the paired t-test of the study’s primary outcome of insomnia severity. Boxplots represent the means and standard deviation of the Insomnia Severity Index (ISI) scores at baseline and after each intervention period. all statistically significant (p=<0.05) differences between intervention periods and baseline are pointed out in this figure.

#### Changes in objective and subjective sleep measures and DLMO

In general, no evidence was found that sleep measures changed between the two intervention periods with the exception of light sleep and sleep timing (**table 2**). Participants had 2.38 percent point (95%CI: -0.22, -2.26) less light sleep during the active morning period compared to the active evening period. Moreover, the percentage of light sleep was 3.1 percent point higher during the active evening period compared to the sedentary period (95%CI -5.83, -0.31) (**supplementary table 3**). The average sleep onset time was 0:30 during the active evening period and 0:13 during the active morning period (delta: 17 minutes, 95%CI: -29, -5). The midpoint of sleep was 3:40 during the active morning period and 4:00 during the active evening period (delta: 20 minutes, 95%CI: -31, -8). Wake up time was 7:28 during the active evening intervention and 7:05 during the active morning period (delta: 23. 95%CI: -41, -4). No differences were observed in DLMO clock time between the interventions. Suggestively, the phase angle between DLMO and sleep onset was on average 17 minutes (95%CI: -0:55, 0:20) shorter during the active morning period compared to the active evening period.

**Table 2.** Comparison of sleep features between active morning and active evening.

|  | Active evening | Active morning | Paired delta | 95% CI |  | Two sided p-value |
| --- | --- | --- | --- | --- | --- | --- |
| | Mean ± SD<br>Median (IQR) | Mean ± SD<br>Median (IQR) | | Lower | Upper | |
| **Sleep duration (min)** | 417 ± 64 | 412 ± 85 | -5.38 | -26.33 | 15.58 | 0.60 |
| **Sleep latency (%min)** | 27.4 (23.1-35.6) | 25.1 (20.1-31.7) | -1.09 | -15.60 | 6.29 | 0.44 |
| **Sleep efficiency (%)** | 86.1 (78.5-89) | 83.6 (76.1-89.2) | -0.57 | -3.98 | 2.18 | 0.59 |
| **Number of awakening at night (n)** | 3.7 ± 1.8 | 3.7 ± 2 | -0.09 | 0.31 | -0.45 | 0.65 |
| **Awake duration (%min)** | 33 (20.1-90.1) | 36.6 (20.7-71.3) | -1.41 | -14.34 | 20.63 | 0.59 |
| **Light sleep (%)** | 62.3 ± 13.8 | 60 ± 14.5 | -2.38 | -0.22 | -2.26 | 0.03 |
| **Deep sleep (%)** | 22.9 ± 11.5 | 24 ± 12.5 | 1.15 | -0.72 | 3.03 | 0.22 |
| **REM sleep (%)** | 14.8 ± 6.6 | 16 ± 7.3 | 1.22 | -0.35 | 2.80 | 0.12 |
| **Phase angle (h:mm)** | 2:33 ± 1:32 | 2:19 ± 1:17 | -0:17 | -0:55 | 0:20 | 0.36 |
| **DLMO (h:mm)** | 21:43 ± 1:12 | 21:39 ± 1:16 | -0:04 | -0:41 | 0:34 | 0.85 |
| **Sleep onset time (h:mm)** | 0:30 ± 1:01 | 0:13 ± 0:59 | -0:18 | -0:29 | -0:05 | 0.006 |
| **Midpoint of sleep (h:mm)** | 3:55 ± 0:38 | 3:39 ± 0:40 | -0:20 | -0:24 | -0:08 | <0.001 |
| **Wake up time (h:mm)** | 7:28 ± 0:47 | 7:05 ± 0:56 | -0:23 | -0:41 | -0:04 | 0.017 |
| **Satisfaction with sleep (0-100)** | 59.2 ± 21 | 55.1 ± 18.8 | -4.03 | -10.40 | 2.35 | 0.21 |
| **Rested feeling (0-100)** | 57.3 ± 22.9 | 53.8 ± 18 | -3.54 | -8.97 | 1.88 | 0.19 |

Min, minute; REM, Rapid Eye Movement; DLMO, Dim Light Melatonin Onset. Table shows means and medians from all sleep parameters during the active morning period and the active evening period. The results from the paired t-test between the active morning and active evening period for all sleep outcomes are also included. Results are shown as mean difference between the groups accompanied by a 95 % confidence interval and a p-value. A p-value equal to or lower than 0.05 was considered a significant difference. Sleep latency, sleep efficiency and awake duration were log transformed and retransformed into a percentage difference.
Sleep latency was excluded in the variable awake duration.

**Table 3.** Sleep variables per benefit group.

|  | **ISI benefit from active morning (n=8)** |  |  | **ISI benefit from active evening (n=9)** |  |  |
| --- | --- | --- | --- | --- | --- | --- |
|  | *Mean active evening* | *Mean active morning* | *Paired delta* | *Mean active evening* | *Mean active morning* | *Paired delta* |
| **ISI score (0-24)** | 14.9 ± 4.1 | 10.3 ± 4.5 | - 4.6 | 9.3 ± 3 | 12.3 ± 2.9 | + 3 |
| **Sleep duration (min)** | 420 ± 94 | 431 ± 125 | + 11 | 423 ± 59 | 405 ± 78 | - 19 |
| **Sleep efficiency (%)** | 88.1 (67.3-90.1) | 88.4 (70.6-91) | - 0.50 | 86.7 (81.9-88.7) | 85.4 (79.5-88.4) | - 2.15 |
| **Sleep latency (min)** | 32 (24-39) | 25 (21-47) | + 1 | 28 (25-36) | 28 (22-38) | + 0.21 |
| **No awakenings at night (No)** | 3.5 ± 2.3 | 3.4 ± 2.6 | - 0.12 | 4.2 ± 1.9 | 3.5 ± 2 | - 0.72 |
| **Awake duration (min)** | 25 (16-142) | 21.4 (15-116) | - 13 | 28 (21-43) | 29.7 (21-56) | - 0.87 |
| **Light sleep (%)** | 64.2 ± 14.6 | 60.6 ± 17.7 | - 3.64 | 60.4 ± 12.4 | 58 ± 10.3 | - 2.33 |
| **Deep sleep (%)** | 20.5 ± 11.8 | 22.2 ± 13.7 | + 1.74 | 25.5 ± 13 | 26.2 ± 13.2 | + 0.72 |
| **REM sleep (%)** | 15.4 ± 4.3 | 17.3 ± 7.6 | + 1.90 | 14.2 ± 5.5 | 15.8 ± 5.9 | + 1.62 |
| **Phase angle (h:mm)** | 2:57 ± 2:23 | 2:26 ± 1:16 | - 54.67 | 2:39 ± 0:56 | 2:38 ± 1:19 | + 10.14 |
| **DLMO (h:mm)** | 22:07 ± 1:25 | 21:50 ± 0:53 | + 0:18 | 21:40 ± 1:11 | 21:12 ± 1:41 | - 0:40 |
| **Sleep onset time (h:mm)** | 1:04 ± 1:34 | 0:27 ± 1:27 | - 0:37 | 0:19 ± 0:36 | 0:02 ± 0:51 | - 0:17 |
| **Midpoint of sleep (h:mm)** | 4:23 ± 0:50 | 4:04 ± 0:54 | - 0:32 | 3:52 ± 0:25 | 3:25 ± 0:28 | - 0:27 |
| **Wake time (h:mm)** | 8:05 ± 0:49 | 7:39 ± 1:11 | - 0:26 | 7:23 ± 0:40 | 6:46 ± 0:44 | - 0:36 |
| **Satisfaction with sleep (0-100)** | 52.5 ± 19.4 | 61 ± 17.7 | + 8.48 | 73 ± 15.4 | 62.1 ± 12.9 | - 10.89 |
| **Rested feeling (0-100)** | 49.4 ± 19.9 | 51.2 ± 15.7 | + 1.79 | 72.3 ± 18.9 | 61.3 ± 15.6 | - 10.94 |

ISI, Insomnia Severity Index; min, minute; DLMO, Dim Light Melatonin Onset. Results are shown as mean ± standard deviation or as median (interquartile range) for non-normal data. The subgroup ‘Benefit from active morning’ contains participants who scored at least 2 points lower on the insomnia severity index after the active morning period compared to the active evening period. The subgroup ‘Benefit from active evening’ contains participants who scored at least 2 points lower on the insomnia severity index after the active evening period compared to the active morning period. The subgroup of participants who’s ISI score did not change 2 point or more was approximately one third (n=10) and is not included in this table. Delta’s were calculated as active evening – active morning for the benefit from active morning group and vice versa for the other group to enhance interpretation.

#### ISI benefit groups

Our data on group level showed no differences between morning vs evening physical activity in most sleep outcomes. However, when inspecting the data on an individual level, relevant changes in both directions in ISI scores can be identified. About one third of the participants (referred to as active morning benefit n=8) showed clinical improvement of ISI scores (change in ISI of at least 2) after the active morning period compared to the active evening period (ISI score active morning 10.3 ± 4.5, delta= -4.6), one third (referred to as active evening benefit, n=9) of participants showed improved ISI scores after the active evening period compared to the active morning period (ISI score active evening 9.3 ± 3, delta= -3) and one third of participants did not have a clinically relevant difference between the active morning and active evening period (ISI score delta <2) (**table 3**). Interestingly, evening chronotypes were significantly (Fishers exact test *p*=0.05) overrepresented among the active morning benefit group (**supplementary figure 1**). During the active morning intervention, the sleep timing of these participants was advanced by approximately 30 minutes (midpoint of sleep 4:04 ± 0:54, delta: - 32 minutes), their phase angle between DLMO and sleep onset was 54 minutes shorter, their sleep duration was 11 minutes longer, and their satisfaction with sleep increased by 8.48 points (mean score: 61 ± 17.7) during the active morning period compared to the active evening period. In the active evening benefit group (consisting mostly of early and intermediate chronotypes, see supp fig 1), subjective sleep improved by approximately 11 points during the active evening period compared to the active morning period (mean score satisfaction with sleep: 73 ± 15.4, delta: 10.89; mean score rested feeling: 72.3 ± 18.9, delta: 10.94). In this group, sleep timing also advanced by approximately 30 minutes during the active morning period compared to the active evening period (midpoint of sleep: 3:25 ± 0:28, delta: 0:27).

## Discussion

The present study is among the first randomized cross-over trials examining how physical activity timing affects insomnia and sleep quality in older adults with sleep issues. Among study completers, we found no clear differences in insomnia severity or subjective and objective sleep quality across timing regimens. However, physical activity timing did influence sleep timing: morning activity led to earlier sleep and wake times than evening activity. This suggests timed physical activity may help adjust sleep timing. Additionally, our observation that evening chronotypes were overrepresented among those whose insomnia improved after morning activity implies chronotype may moderate intervention effects.

Our finding that sleep midpoint was earlier during morning activity aligns with prior studies.(22, 23, 37) A recent randomized controlled trial with 59 men showed that 12 weeks of morning moderate-intensity activity advanced DLMO and reduced sleep latency.(37) A 2004 cross-over trial found earlier sleep timing with morning activity but no changes in other sleep parameters.(38) Sleep timing has been associated with health outcomes. A systematic review linked later sleep timing to poor mental health, cognitive decline, high adiposity, cardiometabolic risk, and decreased bone health in adults.(39) Some Mendelian randomization studies suggest a causal role of a later midpoint of sleep in increasing prostate cancer risk and worse mental health outcomes.(40, 41) Whether these effects stem from sleep timing itself or from social jetlag in late sleepers (42) remains unclear. In our retired population, social jetlag was minimal. Therefore, whether the changes in sleep timing induced by the timing of activities is associated to health outcomes in this population, remains to be investigated.

Despite earlier sleep timing after morning activity, we saw no DLMO changes, suggesting no shift in circadian phase. Although the role of physical activity as a non-photic Zeitgeber is well-established in controlled laboratory studies(13, 23) and multiple real world settings(37, 43), our study did not recapitulate this effect in real-world settings. This discrepancy may be due to the short intervention duration, participant characteristics, or activity intensity. These findings highlight the need for translational research to bridge lab results and real-world interventions affecting circadian timing.

We found no differences in most subjective or objective sleep parameters based on activity timing. There have been a few similar intervention studies performed in the past. A systematic review of nine studies found morning exercise improved sleep quality measured by polysomnography (PSG) and the Pittsburg Sleep Quality Index (PSQI).(44) Another study showed morning light-intensity activity improved total sleep time and wake after sleep onset in older adults without insomnia, while adding low-intensity stepping improved sleep latency and satisfaction with larger effect sizes.(45) A pilot study with 12 older adults without sleep problems found no timing differences and suggested a ceiling effect may have limited results.(38) In this study, participants had a relatively low ISI score (on average subclinical) which could also have influenced the effect sizes. Another systematic review(46) deemed these studies low quality due to selection bias, lack of controls or randomization, and small samples. In our study, participants were randomized and due to the cross-over design, participants were their own control. We did however have a drop-out rate of nearly 25% mostly due to personal or commitment issues. Combined with prior quality concerns, this underscores the challenge and complexity of studying activity timing in real-world settings.

We did however detect a clinical difference in ISI of all three intervention periods compared to baseline. Due to practical reasons, there was a period of approximately two weeks between the baseline measurement and the start of the sedentary period. Although participants were free of any intervention-related assessments and assignments, it is likely that enrollment in the study motivated participants to positively change their behavior towards supporting better sleep quality. This participation effect or ‘Hawthorne’ effect has considerable implications for the generalizability of this study and should be taken into account when designing future studies.(47)

Although group-level effects were absent, individual responses varied considerably. Several participants showed meaningful ISI score changes with corresponding sleep parameter shifts, suggesting differential responses to morning vs. evening activity. Notably, late chronotypes were overrepresented among those who benefitted from morning activity. In this group, sleep timing advanced by ∼30 minutes, and the phase angle for DLMO relative to sleep onset was 55 minutes shorter during morning activity compared to evening activity. This is surprising given prior findings that insomnia patients typically have shorter phase angles than controls.(36) Although exploratory and underpowered for subgroup analysis, these trends suggest that physical activity timing effects vary individually and may be influenced by chronotype. In combination with a previous study showing a chronotype-dependent effects of exercise timing(22) this variation warrants further investigation in larger studies to assess potential for personalized interventions.

This study has several strengths which include it’s study design, rich and continuous data collection and the embedding of the physical activity intervention into activities of daily living of healthy older adults. This study also has several limitations. The most important limitation is the small number of participants. Given that our study was performed in a fixed time frame and all participants underwent the study at the same time and where thus not able to continue inclusion of additional participants, we did not meet the sample size that we defined in our protocol(27) (notably, 40 participants). Furthermore, due to a lack of evidence from previous literature, this initial power calculation was based on the effect size of the absolute difference between physical activity vs. no physical activity. It is very likely that our analyses, especially for the secondary outcomes were therefore underpowered. Due to our small sample size, we also chose not to perform a multiple testing correction. This study could only be conducted in a short time window to prevent between-person seasonal effects and the confounding effect of the transition to daylight saving time in spring which has limited the accessibility to participate. Since the study was conducted from April to June which is a popular holiday-period for retired people in the Netherlands, a total of 37 of the screened people were excluded due to planning issues. Intervention studies with larger study populations (for example, a study with multiple sites) are needed to further test the current hypothesis. We recommend researchers to carefully consider all practical issues of conducting an intervention study about behavioral outcomes like physical activity timing.

In conclusion, this randomized cross-over study suggests that morning physical activity can cause an advance in sleep timing in older adults with sleep problems compared to evening activity. We did not find relevant differences in other objective and subjective sleep parameters between morning and evening physical activity, nor in the phase of the biological clock as measured by DLMO. We did observe considerable variability in individual responses and that late chronotypes benefitted most from morning physical activity with regards to sleep timing and quality in exploratory analyses. With the mounting evidence on the importance of circadian timing of physical activity, we emphasize the need of intervention studies with large study populations and stratifications by chronotype to support the conceptualization of optimal circadian timing of physical activity to promote better sleep and health.

## Data Availability

In the future, the datasets used and/or analyzed during the current study available from the corresponding author on reasonable request. Trial results will be shared or published on multiple websites within the target group and in open access journal as scientific reporting.

## Acknowledgements

We would like to thank the participants of the ON TIME study for their large commitment and contribution to the study.

## Funding

The present work was supported by a grant from the Dutch Research Council (NWO, Dutch National Research Agenda, Research along routes by consortia, 2021–2026, BioClock: the circadian clock in modern society, NWA.1292.19.077; to LK, DvH and DvB). The funding body for this study was not involved in the design of the study nor in the collection, analysis and interpretation of the data or in writing the manuscript.

## Preprint repositories

The abstract of this study was presented as a poster during the annual meeting of the Society for Light, Rhythms, and Circadian Health (SLRCH) in Boston, Massachusetts, United States of America in June 2025.

## Notes

### Competing Interest Statement

The authors have declared no competing interest.

### Clinical Trial

NCT06613958

### Clinical Protocols

https://doi.org/10.1186/s13063-024-08310-7

### Author Declarations

Ethics committee/IRB of Leiden University Medical Center (the Medical Research Ethics Committee of Leiden, The Hague, Delft) name gave ethical approval for this work Statement on approval To whom it may concern, The accredited medical research ethics committee (MREC) Leiden Den Haag Delft, has reviewed research protocol "ON TIME study: Older adults exercising on time" (NL82335.058.22), MREC registration number P22.095 and gave its approval on 6/8/2023. The MRECs decision is based on the documents as stated in appendix 1 of the original Dutch Approval Document. Yours sincerely On behalf of the MREC Leiden Den Haag Delft Mw. Drs. M.Y. Kapteijn secretary

