## Supplementary figure 1 for "The effect of physical activity timing on insomnia and sleep quality: a randomized cross-over trial in older adults"

**Supplementary figure 1.** Chronotype distribution in the benefit groups

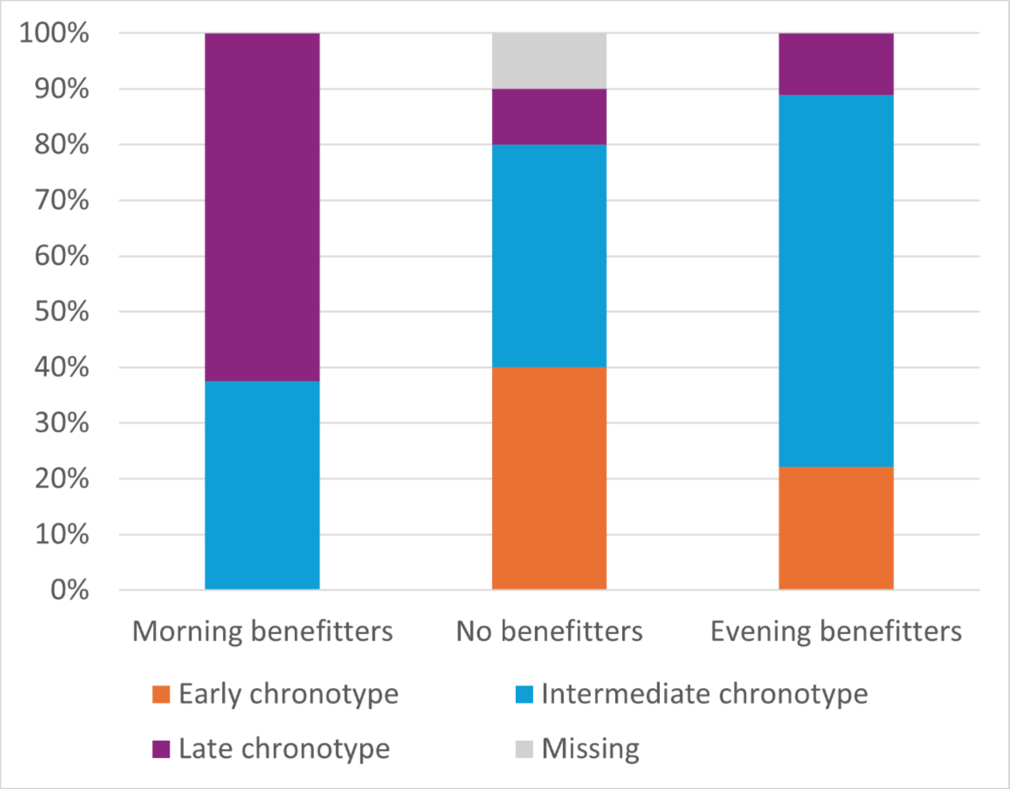


Distribution of chronotypes among the benefit groups. Chronotype was derived from data from the Munich Chronotype Questionnaire (MCTQ). Chronotype groups were based the MSF_sc (sleep-corrected local time of mid-sleep); 25% of participants with the earliest MSF_sc were considered to be 'Early chronotypes', 25% with the latest were defined as 'Late chronotype' and the middle 50% were defined as 'Intermediate chronotype'. The subgroup 'Benefit from active morning' (n=8) contains participants who scored at least 2 points lower on the insomnia severity index after the active morning period compared to the active evening period. The subgroup 'Benefit from active evening' (n=9) contains participants who scored at least 2 points lower on the insomnia severity index after the active evening period compared to the active morning period. The subgroup of participants who's ISI score did not change 2 point or more was approximately one third (n=10).
