## Supplementary table 1 for "The effect of physical activity timing on insomnia and sleep quality: a randomized cross-over trial in older adults"

**Supplementary table 1.** In- and exclusion criteria

**Inclusion criteria**

- Aged between 60 and 80 years old
- Retired
- Long lasting (≥ 3 months) sleep problems (≥10 on the ISI)
- Access to and ability to use a smartphone (Android or Apple)

**Exclusion criteria**

- Currently employed or working
- Participation in any sort of fasting regimen (e.g., intermitted fasting or Ramadan)
- Experienced recent (<6 months) adverse life events (e.g., death of partner)
- Abnormal values in glucose metabolism, thyroid, liver or kidney function, or inflammation markers that after examination of the study doctor need immediate attention of a general practitioner or specialist.
- Diagnosed clinical depression
- Clinically diagnosed neurodegenerative diseases (Dementia or Parkinson's disease)
- Diagnosed sleep apnoea
- Diagnosed restless legs syndrome
- Use of beta-adrenergic blocking agents
- Sporadic use of sleep medication or melatonin supplements
- Injuries or other severe physical conditions (such as active arthrosis) that inhibits physical activity
- Travelled across time zones one week prior to start of study
