## Supplementary table 2 for "The effect of physical activity timing on insomnia and sleep quality: a randomized cross-over trial in older adults"

**Supplementary table 2** Paired comparison of Insomnia Severity Index between baseline and/or intervention periods

| | Pair | Delta | 95% CI | | Two sided p-value |
|---|---|---|---|---|---|
| | | | *Lower* | *Upper* | |
| **Baseline** | Sedentary | 1.94 | 0.77 | 3.1 | 0.002 |
| | Active morning | 2.48 | 1.14 | 3.83 | <0.001 |
| | Active evening | 2 | 0.63 | 3.38 | 0.006 |
| **Sedentary** | Active morning | 0.59 | -0.84 | 2.03 | 0.40 |
| | Active evening | 0.11 | -1.06 | 1.28 | 0.85 |
| **Active morning** | Active evening | -0.48 | -2.02 | 1.06 | 0.53 |

Data from the paired t-test analyses for Insomnia Severity Index (ISI). Data is shown as delta between two of the intervention periods (in points scored on the ISI) accompanied by the 95% confidence interval and p-value.
