## Supplementary table 3 for "The effect of physical activity timing on insomnia and sleep quality: a randomized cross-over trial in older adults"

**Supplementary table 3.** Comparison of sleep features between sedentary and active morning or active evening interventions

| | **Sedentary period** | **Paired delta** *morning - sedentary* | **95% CI** | | **Two sided p-value** | **Paired delta** *evening - sedentary* | **95% CI** | | **Two sided p-value** |
|---|---|---|---|---|---|---|---|---|---|
| | *Mean ± SD* | | *Lower* | *Upper* | | | *Lower* | *Upper* | |
| **Sleep duration** (min) | 427 ± 63 | -4.85 | -18 | 27 | 0.66 | -2.81 | -19 | 21 | 0.92 |
| **Sleep latency** (%min) | 25 (18-35) | -0.66 | -14.80 | 14.27 | 0.91 | 0.73 | -15.14 | 6.11 | 0.50 |
| **Sleep efficiency** (%) | 84 (80-89) | 0.04 | -3.46 | 3.15 | 0.96 | 0.35 | -2.74 | 2.18 | 0.81 |
| **Number of awakening at night** (n) | 3.7 ± 1.9 | -0.07 | -0.23 | 0.38 | 0.63 | -0.04 | -0.32 | 0.33 | 0.97 |
| **Awake duration** (%min) | 37 (22-59) | -0.46 | -34.04 | 17.80 | 0.69 | 0.15 | -20.32 | 17.88 | 0.86 |
| **Light sleep** (%) | 59 ± 14 | 0.63 | -36 | 23 | 0.66 | 3.07 | -5.83 | -0.31 | 0.03 |
| **Deep sleep** (%) | 25 ± 12 | -0.60 | -19 | 31 | 0.63 | -1.71 | -0.60 | 42 | 0.13 |
| **REM sleep** (%) | 16 ± 9 | -0.03 | -19 | 20 | 0.98 | -1.36 | -0.84 | 31 | 0.25 |
| **Phase angle** (h:mm) | 2:34 ± 1:29 | -0:14 | -0:13 | 0:41 | 0.29 | -0:06 | -0:40 | 0:51 | 0.80 |
| **DLMO** (h:mm) | 21:41 ± 1:04 | 0:05 | -0:29 | 0:20 | 0.72 | -0:05 | -0:41 | 0:51 | 0.82 |
| **Sleep onset time** (h:mm) | 0:23 ± 1:22 | -0:16 | - 0:07 | 0:39 | 0.17 | -0:01 | -0:19 | 0:22 | 0.91 |
| **Midpoint of sleep** (h:mm) | 3:57 ± 1:27 | -0:19 | -0:06 | 0:43 | 0.13 | -0:02 | -0:26 | 0:26 | 0.89 |
| **Wake time** (h:mm) | 7:30 ± 1:43 | -0:21 | -0:11 | 0:52 | 0.19 | -0:04 | -0:29 | 0:36 | 0.82 |
| **Satisfaction with sleep** (0-100) | 58 ± 20 | -3.26 | -4 | 11 | 0.37 | 0.77 | -7 | 5 | 0.74 |
| **Rested feeling** (0-100) | 57 ± 21 | -2.96 | -4 | 10 | 0.38 | 0.58 | -6 | 6 | 0.99 |

Min, minute; REM, Rapid Eye Movement; DLMO, Dim Light Melatonin Onset. Table shows means and medians from all sleep parameters during the sedentary period and results from the paired t-test between the sedentary intervention and both the active morning and active evening period for all sleep outcomes. Results are shown as mean difference between the groups accompanied by a 95 % confidence interval and a p-value. A p-value equal to or lower than 0.05 was considered a significant difference. Sleep latency, sleep efficiency and awake duration were log transformed and retransformed into a percentage difference.
Sleep latency was excluded in the variable awake duration.
